# Person-specific Characteristics of People with Low Back Pain Moderate the Preferred Movement Pattern within Motor Skill Training and Strength and Flexibility Exercise

**DOI:** 10.1101/2022.02.28.22271619

**Authors:** Quenten L. Hooker, Linda R. van Dillen

**Author notes:** Corresponding author: Linda R. van Dillen, 4444 Forest Park Boulevard, Campus Box 8502, Program in Physical Therapy, Washington University in St. Louis, School of Medicine, St. Louis, MO 63108, United States of America.

## Abstract

**Background:** People with chronic low back pain (LBP) display an altered movement pattern where the lumbar spine moves more readily into its available range of motion relative to other joints when performing a movement. Recently a randomized controlled trial was completed to compare the effects of motor skill training (MST) to strength and flexibility exercise (SFE). MST improved the altered pattern to a greater extent than SFE. However, there was substantial variability in the baseline and the change over time in the pattern. Understanding factors that influence this variability may ultimately be used to better target treatment strategies to the person.

**Objective:** Examine if gender, age, LBP duration, and the movement pattern at baseline moderate the baseline movement pattern and the change over time in the pattern within MST and SFE. *Design:* Secondary analysis of kinematic data from a single-blind, randomized controlled clinical trial.

**Setting:** Institutional

**Patients:** 154 patients with chronic LBP.

**Interventions:** Motor skill training and strength and flexibility exercise.

**Main outcome measures:** lumbar contribution (LC) to total movement.

**Results:** There was not a significant difference in baseline LC between MST and SFE (β=-2.39, CI=[-7.74, 2.96], p=0.38). SFE did not change LC over time (β=-0.11, CI=[-0.47, 0.24], p=0.53). However, there was a significant change over time in LC within MST (β=-2.13, CI=[-2.54, -1.48], p<0.001). Irrespective of treatment group, there was a trend for gender (β=-5.29, CI=[-10.34, 0.30], p=0.05) and age (β=-0.22, CI=[-0.46, 0.00], p=0.05) to moderate baseline LC. Age (β=0.01, CI=[0.00, 0.02], p = 0.04) and baseline LC (β=-0.07, CI=[-0.10, -0.04], p<0.01) were associated with the change over time in LC within MST only.

**Conclusions:** Person-specific characteristics moderate the baseline altered movement pattern within MST and SFE, as well as the change over time in the pattern within MST.

## Introduction

The primary reason people with LBP seek medical treatment is difficulty in performing daily functional activities.^1,2^ Given the importance of function, researchers have investigated aspects of how people with LBP perform functional activities. Numerous differences in movement characteristics have been reported between back healthy controls and people with LBP.^3,4^ However, specific movement characteristics (e.g., movement velocity) often do not explain substantial variance in LBP-related function.^5^ One set of studies in particular has documented that compared to back healthy controls, people with chronic LBP display an altered movement pattern where the lumbar spine moves more readily into its available range of motion compared to other joints (e.g., knee and hip).^6,7^ Previously, the altered pattern has been indexed as the magnitude of early (first 50% of descent) lumbar movement during an activity. This pattern is of particular clinical relevance because across multiple studies the magnitude of altered movement is associated with LBP symptoms and functional limitations.^6,8,9^ Therefore, the altered pattern is prevalent in people with chronic LBP and relevant to the clinical presentation.

Exercise-based treatment can improve the altered movement pattern during functional activities in people with chronic LBP.^10^ For example, motor skill training (MST) uses person-specific, challenging practice to drive learning of new motor skills.^11-13^ The primary goal of MST during LBP-limited functional activities is to replace the long-standing, pain-provoking, altered movement pattern with a pain-free pattern.^10,12,13^ One study documented that one session of MST immediately improved the altered movement pattern during a functional activity test of picking up an object (PUO), and the improved pattern was associated with an immediate reduction in LBP symptoms.^10^ More recently, the short- and long-term effects of MST were compared to strength and flexibility exercise (SFE). After 6 weeks of MST, on average, people with chronic LBP improved their movement pattern.^14^ Specifically, there was a significant decrease in early lumbar excursion and increase in early knee and hip excursion during the PUO test.^14^ The improved pattern was maintained 6 months after treatment.^14^ SFE, however, did not change the movement pattern across the study duration.^14^ Furthermore, the improved movement pattern in MST corresponded with greater improvements in pain and function compared to SFE.^13^ Although on average MST improved the altered movement pattern and SFE did not, there was substantial variability in the baseline and the change over time in the pattern within both treatment groups. This variability suggests that some people with LBP improved the pattern to a greater extent than others. In order to better refine MST and SFE for people with LBP, it is critical to understand the variability from one person to another in the (1) altered movement pattern at baseline and (2) change over time in the pattern.

One reason for the substantial variability in the pattern at baseline and change over time is that person-specific characteristics (e.g., gender, age, etc.) may influence the pattern. Based on prior reports, there are many person-specific characteristics that have the potential to influence the pattern. First, prior studies suggest there are differences in movement patterns between men and women.^15-20^ Thus, there may be characteristics specific to gender that influence the altered movement pattern at baseline and the change over time in the pattern.^21,22^ Second, previous work suggests a person’s age is associated with movement characteristics, such as spinal range of motion,^23,24^ as well as the clinical course of LBP.^25,26^ Therefore, age could influence the baseline pattern, as well as the effectiveness of the exercise-based intervention. Third, duration of LBP is frequently reported as a prognostic factor of LBP recovery.^26^ Given the previously reported relationship of the altered movement pattern and clinical outcomes,^6,8,9^ duration of LBP may explain the baseline and change over time in the pattern. Lastly, in prior work the magnitude of the altered movement pattern at baseline was associated with the amount of improvement after one session of MST.^10^ Additional research is needed to understand if a relationship also exists between the magnitude of baseline altered movement pattern and the change over time. Understanding the relationship of these person-specific characteristics to the baseline pattern and change over time pattern may ultimately be used to better target treatment strategies to the person and improve the effectiveness and efficiency of care.

The purpose of this study was to examine if person-specific characteristics of gender, age, duration of LBP, and baseline altered movement pattern are associated with both the baseline and the change over time in the altered movement pattern within the MST and SFE treatment groups. We hypothesized that the above-mentioned person-specific characteristics would moderate the baseline pattern and change over time in the pattern.

## Methods

### Participants

This is a planned secondary analysis of kinematic data from a single-blind, prospective, randomized controlled clinical trial (Clinicaltrials.gov: NCT02027623). 154 people with chronic^27,28^ non-specific^29^ LBP were recruited by way of word of mouth, ads through local media, clinics in the region and flyers placed in the community. Inclusion criteria for the study included 1) 18-60 years of age, 2) chronic^27,28^ LBP for at least 1 year, 3) experienced LBP but not in an acute flare-up^30^, 4) modified Oswestry Disability Questionnaire (MODQ)^31^ score of ≥ 20%, 5) difficulty performing at least 3 activities, 6) ability to stand and walk without assistance and 7) ability to understand and sign a consent form. Participants were excluded if they had a BMI >30, any structural spinal deformity, spinal tumor or infection and symptomatic disc herniation by clinical examination. Additional exclusion criteria can be found on Clinical.Trials.gov (ID #: NCT02027623). This study was approved by the Institutional Review Board and all participants provided written informed consent before enrolling in the clinical trial (IRB ID#: 201205051).

### Data collection

Participants completed laboratory sessions for movement analysis at baseline, immediately post-treatment and 6 months after treatment. For the secondary analysis we used a subset of self-report measures that were completed during the clinical trial to describe the study sample. These included 1) demographic and LBP history questionnaire, 2) MODQ, 3) Numeric Rating Scale (NRS) ^32^ for average and worst LBP symptoms (previous 7 days), 4) Fear-Avoidance Beliefs Questionnaire (FABQ)^33,34^ and 5) Short Form Health Survey (SF-36).^35^ Prior to treatment, a standardized clinical examination was performed by a trained physical therapist to classify the person’s LBP.^36-39^ LBP classification was used to guide the person-specific treatment within MST. Classification was based on the person’s altered movement and alignment patterns of the lumbar region and LBP symptom reports during clinical tests.^40,41^

Reflective markers were placed bilaterally on the trunk, pelvis, and lower extremity, according to previously published proceedures.^6,7^ Participants were instructed to perform three trials of a standardized functional activity test of picking up an object (PUO). Measures were obtained at baseline, post-treatment and 6-months post-treatment. For the PUO test, participants stood with their feet pelvis width apart and were asked to pick up the light weight container with both hands and return to the starting position. The light weight container was placed at a height equal to the fibular head and a distance of 50% of trunk length. No instruction was given as to how to complete the PUO test. Further details of the PUO test can be found in Marich et al.^7,10^

Marker co-ordinate data were collected for both a standing calibration trial and the PUO trials using a three-dimensional motion capture system (Vicon Motion Systems, LTD, Denver, CO) with a sampling rate of 120 Hz. Marker trajectory data were tracked using Nexus 2.7.1 (Vicon Motion Systems, LTD, Denver, CO). Data were further processed using custom algorithms written in Visual 3D (C-Motion Inc., Germantown, MD) and MATLAB (MathWorks Inc., Natick, MA). Marker position data were filtered using a 4^th^ order low-pass Butterworth filter with a cut-off frequency of 3 Hz. The cut-off frequency was based on residual analysis^42^ using similar functional activity movement tests.^6,7^ Markers (e.g., lumbar: T12, L3 and S1) were used to create segment specific coordinate systems to track movement across time. Then joint angles were calculated as the distal coordinate system relative to the proximal coordinate system (e.g., hip = femur segment relative to pelvis segment).^6,7^ The primary variable of interest for this study was the magnitude of altered movement pattern. We used lumbar contribution (LC), which was a ratio of early lumbar movement relative to early total movement, to index the magnitude of the altered movement pattern. Early total movement was defined as the summation of early movement of the lumbar spine, hip, and knee joint. Early was defined as the 1^st^ 50% of movement time for the descent phase of the PUO task.^6,7,10^ The lumbar contribution index was calculated using the following equation: 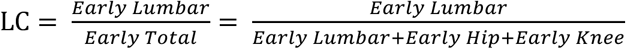. For example, a LC of 0.50 would be interpreted as the lumbar spine contributing 50% of the total movement during the early phase of the PUO test. Therefore, a greater LC represents a greater magnitude of altered movement pattern.

Participants were randomized to either MST or SFE. All participants received 6, 1-hour treatment sessions, scheduled once/week for 6 weeks. Complete details of the treatment conditions can be found on Clinicaltrials.gov (NCT02027623). SFE focused on increasing the strength of all of the trunk muscles and improving all planes of trunk and lower limb flexibility based on ACSM guidelines.^43^ MST involved challenging, person-specific practice to improve pain provoking, altered movement and alignment patterns during the performance of functional activities. The primary objectives of treatment were to train the participant to 1) decrease the amount of early lumbar spine movement related to the person’s LBP classification (e.g., flexion), 2) increase the contribution of movement of other joints (e.g., knees and hips) and 3) avoid end range movements or alignments of the lumbar spine in the specific direction related to the participant’s LBP classification. Physical therapists minimized the extrinsic feedback given to participants during practice. Training focused on problem solving by the participant to learn to perform the activities without increasing LBP symptoms. Both treatment conditions were progressed based on the participant’s ability to perform the exercise or activity independently.^44^

### Data analysis

All analyses were performed using R v3.5.3.^45-47^ Descriptive statistics were calculated for participant demographics and self-report measures. Two-sample t-tests and chi-square tests of independence were used to test for significant differences (p < 0.05) in characteristics between treatment groups at baseline.

We used hierarchical linear modeling (HLM) to examine the moderating effects of gender, age, LBP duration, and baseline LC on the baseline LC and change over time in LC. HLM is a regression-based approach that represents trajectories of participants over time by modeling responses at multiple levels of measurement.^48^ HLM is a recommended strategy for longitudinal data that 1) differ in measurement intervals, 2) are nested in separate levels of measurement (e.g., time within each participant) and 3) contain outcomes of either continuous or categorical data.^48^ Given our study design and variables fit these criteria HLM is an appropriate analytic technique. Initially, the LC outcomes were modeled over time at level 1 (i.e., baseline, post-treatment, and 6 month follow up). The level 1 analysis estimated each person-specific intercept, linear time component, and non-linear time component (i.e., quadratic). Moderators of the person-specific LC outcome trajectories were modeled at level 2. The moderators were treatment group (TxGroup), gender, age, duration of LBP, and baseline LC. Also at level 2, we included the variability in coefficients from level 1, as a function of TxGroup, person-specific characteristics, and the interactions of TxGroup X each characteristic. Specific contrasts of the level 2 effects were calculated to test for the 1) main effect of time, TxGroup, and each person-specific characteristic and 2) interactions among the three domains of variables.

## Results

### Study sample

The study sample included 154 participants. Twenty-one (13.6%) participants withdrew over the study duration. Participant characteristics, grouped by treatment group, are summarized in Table 1. There were no significant differences between MST and SFE for any participant characteristic at baseline (Table 1).

**Table 1:** Mean +/- SD for participant characteristics at baseline by treatment group

|  | **Strength and**<br>**Flexibility Exercise**<br>**(n = 77)** | **Motor Skill**<br>**Training**<br>**(n = 77)** | **p value** |
| --- | --- | --- | --- |
| Age (years) | 42.6 ± 11.7 | 42.5 ± 11.8 | 0.94 |
| Female, no. (%) | 52 (68) | 43 (56) | 0.14 |
| BMI (kg/m<sup>2</sup>) | 25.4 ± 3.2 | 26.1 ± 3.1 | 0.18 |
| Duration of LBP (years) | 10.9 ± 9.3 | 9.7 ± 7.9 | 0.39 |
| LBP Medication Use no. (%) | 48 (62.3) | 45 (58.4) | 0.63 |
| Function (MODQ, 0-100%) † | 32.7 ± 10.2 | 32.5 ± 9.3 | 0.90 |
| Average Pain (NRS, 0-10) ‡ | 4.7 ± 1.8 | 4.7 ± 1.6 | 0.96 |
| Worst Pain (NRS, 0-10) ‡ | 6.3 ± 2.0 | 6.8 ± 1.6 | 0.09 |
| Fear-Work (FABQ-W, 0-42) § | 11.0 ± 8.3 | 11.7 ± 8.9 | 0.62 |
| Fear-Physical (FABQ-P, 0-24) § | 14.9 ± 6.1 | 14.1 ± 5.1 | 0.35 |
| SF-36 Physical ¶ | 42.9 ± 6.8 | 40.7 ± 6.9 | 0.05 |
| SF-36 Mental ¶ | 48.8 ± 11.5 | 52.1 ± 9.2 | 0.05 |

† modified Oswestry Disability Questionnaire scores range between 0% (no LBP-related functional limitation) and 100% (max limitation)
‡ Patient report of average or worst pain in the prior 7 days on verbal numeric pain rating scale between 0 (no pain) and 10 (pain as bad as can be)
§ Fear-Avoidance Beliefs Questionnaire physical activity subscale score ranges from 0-24 and work subscale score ranges from 0-42 with higher scores indicating higher fear-avoidance
¶ 36-Item Short Form Health Survey (SF-36) Physical and Mental Component summary scores are scaled and normalized to have a mean of 50 and standard deviation of 10 in the normal 1998 US population

### Treatment Effects

There was not a significant difference in baseline LC between MST and SFE (β = -2.39, SE =2.73, p = 0.38). Within SFE there was not a significant linear slope (β = -0.11, SE = .18, p = 0.53) or quadratic component (β = 0.001, SE = 0.005, p = 0.86). The specific treatment group comparisons in the change over time in LC indicate that there was a significant difference in the linear slope and curvilinearity (ps < 0.05, Table 2). Within MST there was a significant overall linear slope (β = -2.13, SE = 0.20, p < 0.01) and quadratic component (β = 0.05, SE = .01, p < 0.01). The effects within MST were a result of a decrease in LC during the treatment phase, plateauing after the completion of treatment, and regressing at 6 months (β = 1.04, SE = .18, p < 0.01).

**Table 2:** Results of hierarchical linear modeling analyses of lumbar contribution over time at baseline, post-treatment and 6 month follow up

| **Baseline** |  |  |  |
| --- | --- | --- | --- |
| *Fixed effect* | *Beta coefficient* | *CI* | *p-value* |
| Intercept † |  |  |  |
| TxGroup | -2.385 | (-7.74, 2.97) | 0.384 |
| Gender | -5.288 | (-10.58, 0.01) | 0.052 |
| Age | -0.224 | (-0.45, 0.01) | 0.056 |
| LBP Duration | -0.187 | (-0.49, 0.12) | 0.241 |
| Linear ‡, §, ¶ |  |  |  |
| TxGroup | -2.012 | (-2.55, -1.48) | < 0.001 |
| Gender | 0.056 | (-0.11, 0.23) | 0.511 |
| Age | -0.004 | (-0.01, 0.01) | 0.261 |
| LBP Duration | 0.004 | (-0.01, 0.02) | 0.391 |
| Baseline LC | -0.013 | (-0.03, 0.01) | 0.197 |
| TxGroup X Gender | -0.177 | (-0.41, 0.06) | 0.157 |
| TxGroup X Age | 0.015 | (0.01, 0.02) | 0.003 |
| TxGroup X LBP Duration | -0.010 | (-0.02, 0.01) | 0.155 |
| TxGroup X Baseline LC | -0.058 | (-0.08, -0.03) | < 0.001 |
| Quadratic §, ¶, ¥ |  |  |  |
| TxGroup | 0.049 | (0.04, 0.06) | < 0.001 |
| Baseline LC | 0.001 | (-0.01, 0.01) | 0.302 |
| TxGroup X Baseline LC | 0.002 | (0.00, 0.02) | < 0.001 |

| **Post-Treatment** |  |  |  |
| --- | --- | --- | --- |
| *Fixed effect* | *Beta coefficient* | *CI* | *p-value* |
| Intercept † |  |  |  |
| TxGroup | -16.154 | (-19.54, -12.76) | < 0.001 |
| Gender | -4.973 | (-8.52, -1.42) | 0.036 |
| Age | -0.061 | (-0.22, 0.09) | 0.188 |
| LBP Duration | -0.129 | (-0.34, 0.08) | 0.382 |
| Linear ‡, §, ¶ |  |  |  |
| TxGroup | -1.112 | (-1.43, -0.79) | < 0.001 |
| Gender | -0.045 | (-0.24, 0.15) | 0.655 |
| Age | -0.007 | (-0.02, 0.01) | 0.093 |
| LBP Duration | 0.001 | (-0.01, 0.01) | 0.881 |
| Baseline LC | -0.060 | (-0.07, -0.05) | <0.001 |
| TxGroup X Gender | -0.161 | (-0.43, 0.11) | 0.241 |
| TxGroup X Age | 0.014 | (0.00, 0.03) | 0.017 |
| TxGroup X LBP Duration | -0.007 | (-0.02, 0.01) | 0.393 |
| TxGroup X LC | -0.014 | (-0.03, 0.00) | 0.023 |
| Quadratic §, ¶, ¥ |  |  |  |
| TxGroup | 0.047 | (0.04, 0.06) | < 0.001 |
| Baseline LC | 0.002 | (0.00, 0.01) | < 0.001 |
| TxGroup X Baseline LC | 0.001 | (0.00, 0.01) | 0.013 |

| 6-month follow up |  |  |  |
| --- | --- | --- | --- |
| *Fixed effect* | *Beta coefficient* | *SE* | *p-value* |
| Intercept † |  |  |  |
| TxGroup | -17.961 | (-23.12, -12.80) | < 0.001 |
| Gender | -6.281 | (-11.74, -0.82) | 0.026 |
| Age | -0.056 | (-0.28, 0.17) | 0.613 |
| LBP Duration | -0.145 | (-0.46, 0.17) | 0.364 |
| Linear ‡, §, ¶ |  |  |  |
| TxGroup | 1.470 | (1.05, 1.89) | < 0.001 |
| Gender | -0.088 | (-0.27, 0.09) | 0.341 |
| Age | -0.003 | (-0.01, 0.01) | 0.475 |
| LBP Duration | -0.001 | (-0.01, 0.01) | 0.939 |
| Baseline LC | -0.005 | (-0.02, 0.01) | 0.567 |
| TxGroup X Gender | 0.103 | (-0.06, 0.27) | 0.222 |
| TxGroup X Age | -0.002 | (-0.01, 0.01) | 0.635 |
| TxGroup X LBP Duration | -0.002 | (-0.01, 0.01) | 0.695 |
| TxGroup X Baseline LC | 0.048 | (0.02, 0.07) | < 0.001 |
| Quadratic §, ¶, ¥ |  |  |  |
| TxGroup | 0.052 | (0.04, 0.06) | < 0.001 |
| Baseline LC | 0.001 | (-0.01, 0.01) | 0.198 |
| TxGroup X Baseline LC | 0.002 | (0.00, 0.01) | 0.011 |

TxGroup represents the difference in lumbar contribution (LC) between strength and flexibility exercise and motor skill training.
† Gender, Age and LBP Duration are differences in the grand means, irrespective of TxGroup
‡ Linear slope at the centered time point
§ Main effects are within strength and flexibility exercise
¶ Interaction effects represent the difference between strength and flexibility exercise and motor skill training.
¥ Quadratic component at the centered time point

### Person-Specific Effects

#### Gender

Irrespective of treatment group there was a trend for a significant difference in baseline LC between men and women (β = -5.29, SE = 2.69, p = 0.05). Specifically, men had a greater LC than women across all time points in the study (Figure 1). There was not a significant difference in the linear change over time in LC between men and women within SFE or MST (Table 2). *Age:* Irrespective of treatment group there was a trend for a relationship of age and LC at baseline (β = -0.22, SE = 0.12 p = 0.06). This corresponds to a 0.22 unit decrease in LC for every additional 1 year of age; older people tend to have a smaller baseline LC. Within SFE there was no significant moderating effect of age on the change over time in LC. Alternatively within MST, age moderated the linear change over time (β = 0.01, SE = .004, p < 0.01). Specifically older participants have a flatter (less negative) slope than younger participants (Figure 2). *LBP Duration:* Across MST and SFE there was not a significant relationship of duration of LBP and baseline LC (β = -0.19, SE = .16, p = 0.24) or the linear change over time in LC (Table 2). *Baseline LC:* Within SFE there was no significant relationship of baseline LC and the linear change over*\* time in LC (Table 2). However, in MST there was a significant relationship of baseline LC and the linear change over time in LC (β = -.07, SE = .01, p < 0.01). People within MST with a greater baseline LC had a more negative slope than those with less baseline LC (Figure 1).

**Figure 1.**
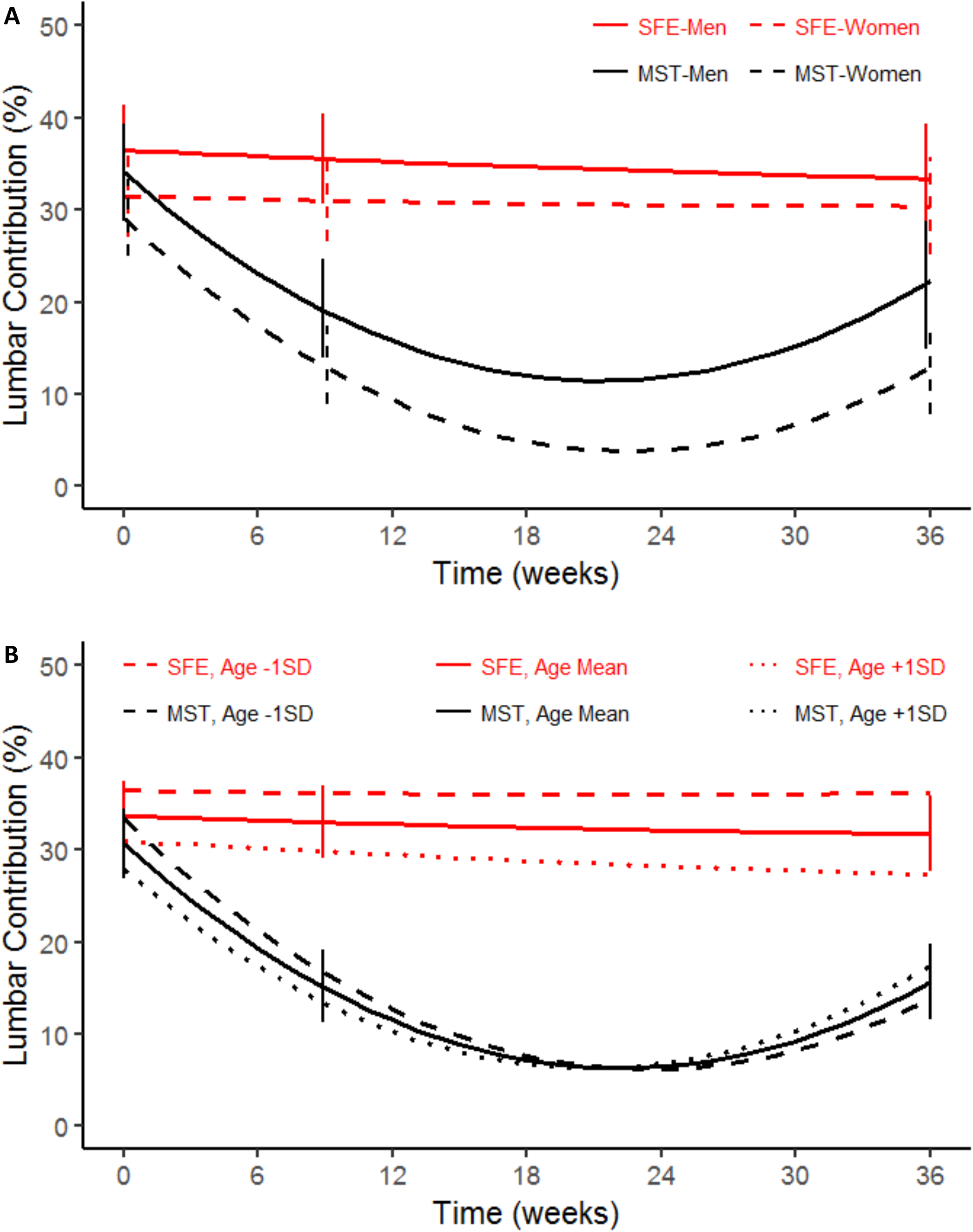
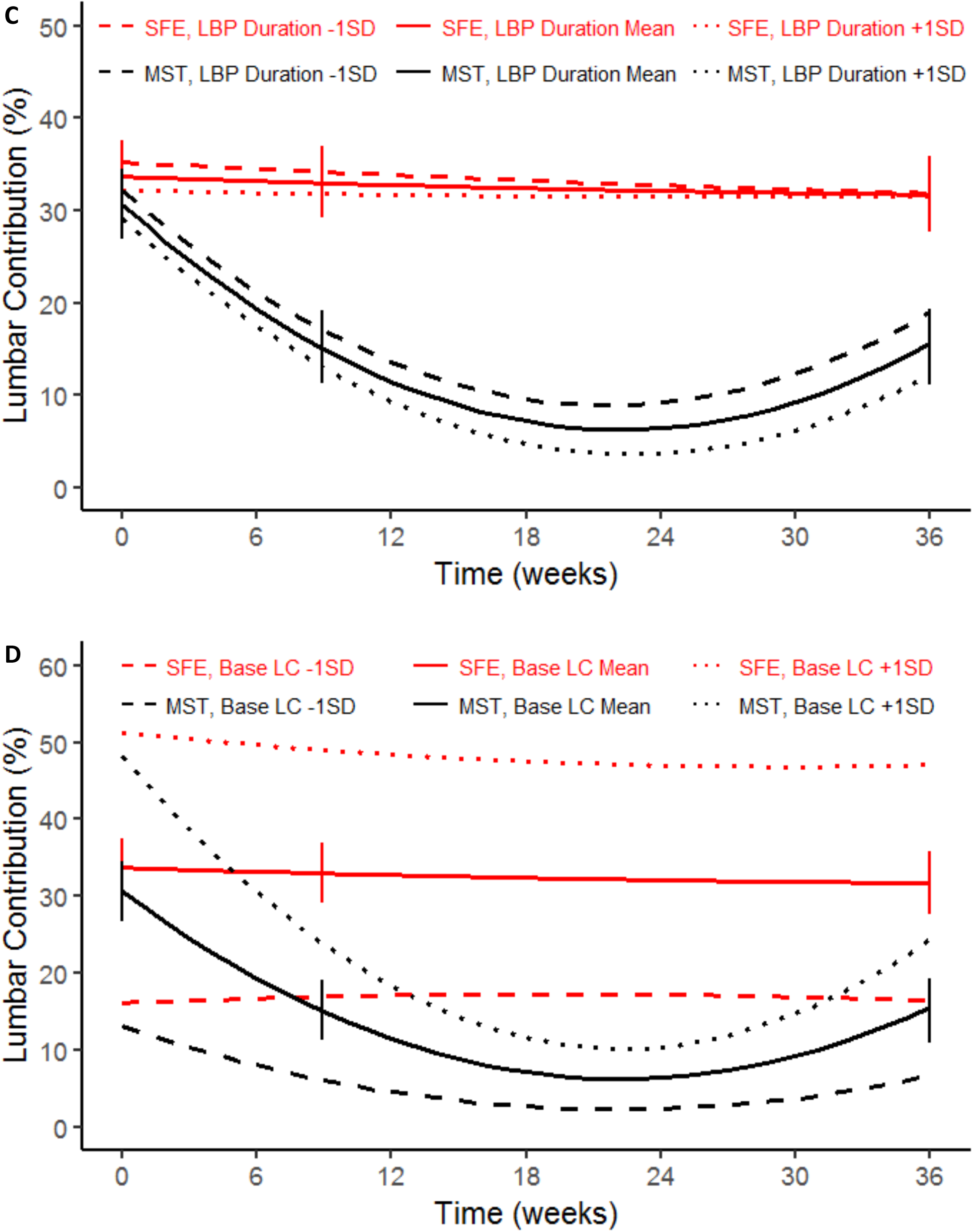
Predicted values based on hierarchical linear modeling analysis of lumbar contribution (LC) for motor skill training (MST) and strength and flexibility exercise (SFE). Confidence intervals are displayed at the three centering locations of baseline, post-treatment and 6 months post-treatment. Person-specific moderating effects are displayed for **1A**. Gender, **1B**. Age, **1C**. Duration of LBP and **1D**. Baseline lumbar contribution (LC_base).

## Discussion

The purpose of this study was to examine if the person-specific characteristics of gender, age, duration of LBP, and magnitude of baseline LC were associated with the baseline pattern and change over time in the pattern within MST and SFE. Our hypothesis that person-specific characteristics would moderate the altered movement pattern was partially supported. First, there was a trend for differences between men and women in baseline LC, but gender did not moderate the change over time in LC within either treatment group. Second, there was a trend for a relationship of age and the baseline LC. However, age only significantly moderated the change over time in LC within MST. Third, duration of LBP was not associated with the baseline or change over time in the altered movement pattern. Finally, baseline LC was associated with the change over time in LC only within MST. Therefore, these data support that person-specific characteristics are relevant to the magnitude of the altered movement pattern at baseline and the change over time in the pattern in MST.

Irrespective of treatment group, there was a trend for a significant difference in baseline LC between men and women. Specifically, men had 5.3% greater LC than women. The modeled differences in LC between men and women were observed consistently across the study duration (ps = 0.03 to 0.05); men moved more in their lumbar spine early relative to other joints compared to women. There also was no difference in the trajectory of change over time in LC between men and women within MST and SFE. Similar to prior research, these data suggest men and women perform functional activity tests differently.^19,20,22^ However, if given the appropriate training to directly improve the altered movement pattern, men and women can both improve to a similar extent. Therefore, a clinician may expect men and women to perform a functional activity test differently, but both genders have a similar capacity to improve the altered movement pattern.^22^

There was a trend for age and baseline lumbar contribution to be associated. Specifically, for people who are 1 standard deviation older (mean + 1SD = 54.3 years) than the mean (42.6 years) there is a 2.6% reduction in lumbar contribution; the opposite effect holds for people who are younger than the sample mean. Interestingly, age also moderated the change over time in lumbar contribution within MST (Figure 1B). For example, older people had a smaller change over time in LC, compared to younger people. Older people also did not retain the improved pattern as well as younger people. Although these relationships are statistically significant, these findings should be interpreted with caution. Even across multiple decades of age, people within MST improve the magnitude of altered movement pattern during the functional activity test of PUO. Therefore, age has a relatively small effect on baseline and the change over time in LC, and MST improved the altered movement pattern in both younger and older people.

We found that the number of years someone has LBP is not associated with the magnitude of altered movement pattern at baseline or the change in the pattern over time. Previously, a moderate positive relationship (r = 0.39) was reported between LBP duration and change in the altered movement pattern after one session of MST.^10^ Specifically, the prior study reported that the longer the person had LBP the greater improvement in the altered pattern after one session of MST.^10^ One reason for the discrepancy from the current study is that prior work examined the change in the pattern after 1, 20 minute session of MST, as opposed to 6, 1-hour treatment sessions that were once/week for 6 weeks.^10^ Thus, the moderate relationship of duration of LBP and change in the altered movement pattern after one session of MST is not observed over the short-term (i.e., 6 weeks) or long-term (i.e., 6 months). The lack of association of LBP duration and change in the pattern over time is of particular importance for those within the MST group, because these data highlight that repeated sessions of MST improved a long-standing, pain-provoking, altered movement pattern, irrespective of how long the person had LBP.

Baseline LC moderated the change over time in LC within MST. Specifically, people with greater LC in MST initially had a greater improvement (more negative slope) compared to people with less LC (Figure 2D). These data mirror the relationship of baseline early lumbar excursion and the change in early lumbar excursion after one 20-minute session of motor skill training.^29^ The people with the greatest magnitude of altered movement pattern at baseline change the most following MST because they have the greatest potential to change. Furthermore, comparing the trajectories of the change over time for those with greater LC in SFE and MST, people with greater baseline lumbar contribution have a vastly different trajectory. Those with a higher level of altered movement pattern in SFE do not improve the altered pattern; whereas those in MST have a rapid improvement. These data further indicate that if a goal of treatment for people with chronic LBP is to improve the altered movement pattern associated with function, MST is superior to SFE.

Although our findings support that there are person-specific factors that may help refine treatment, this study has limitations. First, these findings are relevant to the altered movement pattern during one functional activity test. Additional research is necessary to determine whether these effects are similar in other functional activities that are limited in people with LBP. Second, we excluded people with a BMI > 30 and specific LBP conditions (e.g., disc herniation). Therefore, these data may not be generalizable to all people with chronic LBP. In addition, other person-specific characteristics not tested in this study should be examined.

## Conclusion

Our findings suggest that person-specific characteristics such as gender, age and baseline lumbar contribution moderate the baseline and change over time in the magnitude of the altered movement pattern. Alternatively, duration of LBP is not associated with the pattern at baseline or the change over time. These person-specific characteristics, in part, explain the variable movement pattern trajectories over time. These findings are of specific importance because understanding factors that influence person-level variability can ultimately be used to better target treatment strategies to the person. Lastly, our findings are clinically relevant because even accounting for the person-specific variability in the movement pattern, MST is superior to SFE in improving the altered movement pattern in people with chronic LBP.

## Data Availability

All data produced in the present study are available upon reasonable request to the authors.

## Acknowledgments

The authors would like to thank Sara Putnam, Sara Francois, Kristen Roles, and Jennifer Jarvis for their assistance with participant recruitment, data collection and data processing.

